# Minimal training of fieldworkers in resource-poor settings for rapid assessment of scabies prevalence: a diagnostic accuracy study in Mopeia, Mozambique

**DOI:** 10.1101/2024.02.09.24302523

**Authors:** Joanna Furnival-Adams, Valeria López, Hansel Mundaca, Amelia Houana, Antonio Macucha, Eldo Elobolobo, Aida Xerinda, Humberto Munguambe, Felisbela Materula, Regina Rabinovich, Francisco Saute, Daniel Engelman, Carlos Chaccour

## Abstract

**Background:** Scabies is endemic in many tropical, resource-poor areas, causing significant morbidity. Our understanding of the true burden of scabies in Africa is very limited, and we lack tools to accurately assess its prevalence, given our reliance on highly qualified doctors and microscopy for diagnosis. The primary objective of this study was to assess the accuracy of minimally trained fieldworker diagnosis of scabies, compared to diagnosis by experienced medical doctors.

**Methodology:** We trained 62 fieldworkers with a minimum of secondary school education in the diagnosis of scabies (categorized as either clinical or suspected), and in terms of severity, based on lesion count. Immediately after the training, we assessed their knowledge based on written assessments. We also assessed the diagnostic accuracy of a sub-sample of the fieldworkers in the field compared to the reference standard (experienced medical doctors). 193 individuals were assessed for scabies. The sensitivity, specificity, PPV and NPV were calculated, as well as agreement (kappa coefficients) between medical doctors and between fieldworkers.

**Results:** The overall median results in written assessments were around 80%. Of the 193 people assessed for scabies, 27% were classified as having scabies according to the reference standard. The sensitivity of fieldworker diagnosis compared to the reference standard was 95% (95%CI 93 -100), and the specificity was 99% (95%CI 99-100). The sensitivity for severe scabies was lower at 64% (95%CI 51-75), and the mean specificity was 100%. Kappa coefficients were 1.00 between medical doctors and 0.95 between fieldworkers.

**Conclusions:** Fieldworkers without medical qualifications were capable of diagnosing scabies to a similar level of accuracy as experienced medical doctors, after a short period of focal training. This may facilitate rapid assessments of scabies prevalence for public health purposes and decisions about MDA implementation in similar settings.

**Author summary:** *What is already known on this topic:* Scabies is a disease that causes intense itchiness which can affect people’s wellbeing and ability to attend work or school, and if left untreated it can lead to further, more severe complications. Typically, scabies is diagnosed by specialist medical doctors using microscopy. However, many countries lack these resources and therefore there is a paucity of data on scabies prevalence.

*What this study adds-:* In our study, we trained people with no previous medical training in the diagnosis of scabies over a few days through presentations and assessments. We demonstrated that they were able to diagnose scabies to a similar degree of accuracy compared to experienced medical doctors.

*How this study might affect research, practice or policy-:* Understanding that minimally trained fieldworkers are able to diagnose scabies to a similar degree of accuracy as medical doctors, may facilitate the collection of data for survey purposes in resource-poor areas, which in turn may promote advocacy for the disease.

## INTRODUCTION

Scabies is an ectoparasitic skin infestation, endemic in many tropical, resource-poor communities (1). Scabies mites cause skin lesions and intense itch that are often associated with secondary bacterial infections (impetigo) that can lead to more severe sequelae, such as severe soft tissue infection, rheumatic fever, post-streptococcal glomerulonephritis and sepsis (2,3). In addition to the morbidity associated with scabies, it also has negative profound psychosocial and economic impacts that can perpetuate poverty. Furthermore, effective treatment can be difficult to access in rural, lower-middle income settings, meaning that episodes of scabies are often long-lasting and recurrent.

Our understanding of the true prevalence of scabies in the African region is extremely limited. A cross-sectional analysis of global scabies prevalence in 2017 reported no available data sources for sub-Saharan Africa and relied on estimates from other continents and the covariate of water source to estimate the burden of scabies in this region (4). A more recent article reported estimates of scabies prevalence in eight African countries, however, the majority of these estimates were based on small-scale studies, and there were no available data from the remaining 46 countries (1). High-quality data collection has been impeded by the limited investment in scabies control programmes and research. Paradoxically, high quality data is needed to encourage investment into scabies disease programmes and to generate data. Therefore, accessible and economic methods of assessing scabies prevalence are needed to further advocate for investment into scabies control programmes.

In sub-Saharan Africa (SSA), the most economically disadvantaged communities often live in rural, difficult-to-reach areas (5). Thus, individuals living with scabies often don’t seek care and therefore, the number of cases seen at health facilities is not a reliable indicator of the true scabies burden in the community (1). Therefore, novel diagnostic approaches and tools for assessing scabies in field settings are needed.

Currently, there are no point-of-care diagnostic tests for scabies that are appropriate for use in field settings (6,7). Accessibility and availability of these tools would improve our ability to estimate population prevalence; and provide information that could be used for control programme decision making. Confirmation of scabies diagnosis in high-income clinical settings typically requires examination by an experienced doctor and microscopic visualisation of mites on skin scrapings. However, these methods are not feasible in field settings as they require specialised equipment and highly skilled workers. The need for improved diagnostic tools for scabies has been highlighted as a priority for global scabies control as part of the World Health Organization (WHO) 2030 road map for neglected tropical diseases (NTDs) (8,9). To accommodate this, the consensus diagnostic criteria for scabies were developed in 2020 by a panel of experts convened by the International Alliance for the Control of Scabies (2020 IACS criteria), with the goal of promoting standardised case definitions of scabies and more comparable data sets (10,11). The 2020 IACS criteria define scabies cases as either confirmed, clinical or suspected based on the level of diagnostic certainty. Typically, medical doctors are responsible for assessing whether someone has scabies. However, task-shifting, whereby tasks typically performed by a doctor are shifted to healthcare professionals with less specialised positions, can improve access to healthcare in resource-poor areas (12). The same strategy may be applied for scabies prevalence surveys. There have been a few studies assessing the accuracy non-clinician healthcare professional for the diagnosis of scabies in the field following the IACS criteria in south pacific and west African settings. One study evaluated the diagnostic accuracy of four, briefly-trained nurses who examined a cohort of 171 school children using clinical criteria (13). The researchers found the nurses to have high levels of sensitivity and specificity for moderate to severe scabies (94% and 74% respectively), but low sensitivity for mild scabies, when compared to the consensus diagnosis of two expert physicians. In another study, two dermatologists trained five nurses, and screened 135 people for scabies, and found 55% sensitivity and 90% specificity, with again higher levels of sensitivity for moderate and severe cases (14). Finally, a study evaluated the use of a modified, simplified version of the IACS 2020 criteria for mapping and rapidly assessing scabies prevalence (15) in very-high prevalence settings. This modified version had a sensitivity of 82% and specificity of 98% compared to diagnosis using of the full 2020 IACS criteria, with no difference in the pooled prevalence estimates using the two methods.

In 2022, a large-scale randomised-controlled trial (BOHEMIA) took place in Mopeia, Mozambique assessing ivermectin mass drug administration (MDA) for malaria vector control and enrolling over 20,000 participants (16). A secondary objective of this clinical trial was to assess the impact of ivermectin MDA on the local prevalence of scabies. The fieldworkers who implemented the MDA had a minimum of secondary school education to grade 10. The resources to screen such a large population using medical doctors and microscopy were not available, and therefore we chose to train a sub-group of the same fieldworkers who implemented the MDA in the diagnosis of scabies. If non-medical workers can accurately identify scabies cases, this may facilitate rapid assessments of scabies prevalence for public health purposes and decisions about MDA implementation in similar settings.

The primary objective of this study was to assess the accuracy of minimally trained fieldworker diagnosis of scabies in terms of positive predictive value (PPV), negative predictive value (NPV), sensitivity and specificity, compared to diagnosis by experienced medical doctors.

## METHODS

### Study design and setting

This study was nested within a trial that took place between March-September 2022 in Mopeia, Mozambique. This district is located in the central north of Mozambique and has a population of around 162,000 (17). To date, there have been no large-scale published studies reporting scabies in Mozambique(18).

### Training procedures

We trained 90 individuals with high-school qualifications for two days. Between 4^th^-7^th^ February 2022, 28 supervisors and 62 fieldworkers were trained in the diagnosis of scabies and infected scabies by JFA, VL and AH. JFA has a background in infectious disease epidemiology and extensive knowledge of scabies pathology. VL and AH are medical doctors with experience working and diagnosing scabies in tropical, scabies-endemic settings. All fieldworkers had completed secondary education up to grade 10, but experience in healthcare settings was not a specified requirement. Fieldworkers were selected from a larger team of study workers based on observed performance in training for other trial activities. The training techniques were based on methods used in previous studies (13,19). Day 1 consisted of 5 hours of presentations and discussion on scabies and infected scabies. Day 2 included a written assessment (assessment 1) followed by a summary and discussion of the answers. The slides and structure of the training was designed and agreed upon amongst the three trainers. The assessment included 50 photo-based case scenarios, where the trainees were required to assess the presence or absence of scabies lesions, quantify scabies lesions and assess the presence or absence of impetigo. The trainees were instructed to feel the skin using gloves in milder cases where there may be fewer than 10 lesions. The ground truth for these photos was based on confirmed diagnoses by experienced medical doctors. A refresher training session took place (for fieldworkers only) on 18^th^ February 2022, 11 days after the first sessions, and included a video demonstration; review of the scabies and impetigo presentations; and assessment practice using role-play. The fieldworkers then repeated the written assessment, using the same case images in a different random order (assessment 2). Throughout the training sessions the trainers asked the fieldworkers questions to reaffirm their knowledge and they were encouraged to ask questions if anything was unclear.

### Validation of the fieldworkers’ scabies assessment against the MDs

An assessment of the diagnostic accuracy of the trained fieldworkers took place during a three-week period in July 2022. Each day, two fieldworkers and the two medical doctors visited households that consented to participate in the study, within clusters that had previously reported high levels of body itch during the enumeration and census visits conducted in preparation for the trial (Figure 1).

**Figure 1.**
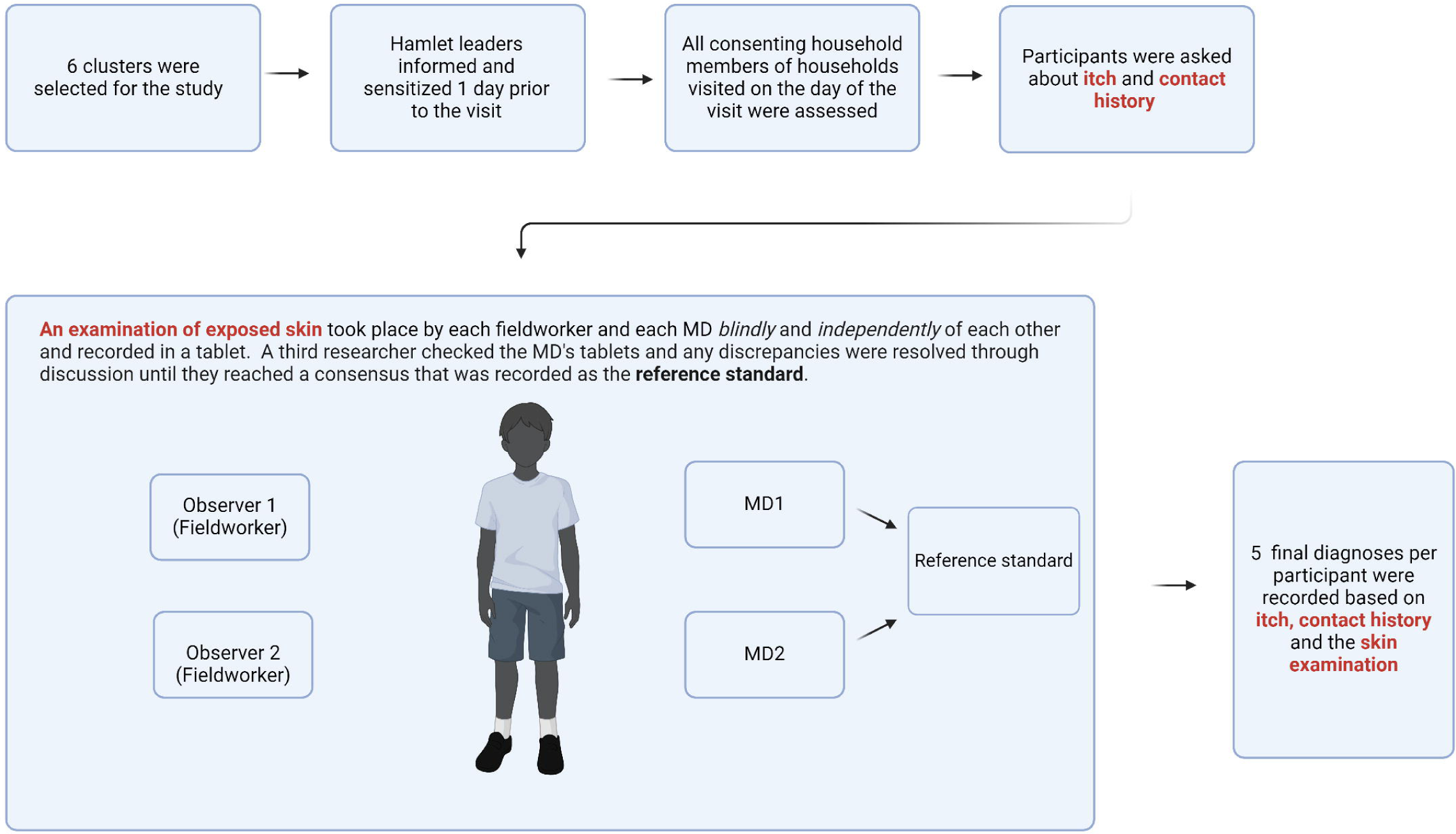

The hamlet leaders of these communities were asked for verbal consent 1 day prior to our visit and made community members aware of the planned visit. All household members who were present on the day of the visit were screened. Nine fieldworkers were randomly selected to take part in the assessment. For every visit, two fieldworkers and the two medical doctors jointly asked participants questions about body itch within the past 24 hours and contact history (body itch amongst other household members within the past 24 hours). Due to the need for all assessors to understand the local languages, it was not possible to conduct this part of the diagnosis independently. Following the questionnaire, the fieldworkers and medical doctors each independently examined the exposed skin of the participant. If lesions were present, the assessors counted the number of lesions and this was used as an indicator of severity, with 1-2 lesions classified as very mild; 3-10 mild; 11-49 moderate and ≥ 50 severe. Following the IACS 2020 diagnostic criteria, all assessors classified participants as either having “clinical scabies”, “suspected scabies”, or “not scabies”.

### Index test and reference standard

After the joint questioning about symptoms and contact history, the fieldworkers were instructed to examine the skin sequentially and without any discussion between them. Then the fieldworkers recorded their results independently and blinded to one another into an electronic questionnaire using a mobile tablet device. The index test was the individual classification of each participant as clinical scabies, suspected scabies, or not scabies by the field workers based on the skin assessment, combined with the participants’ response to the joint questioning.

The two medical doctors examined each participant independently and blinded to one another and entered their observations in the tablet. They then shared their results privately with another researcher (JFA) and, if there were any disagreements, they discussed the case until they reached a consensus. The clinician’s joint assessment based on the history of body itch at personal and household levels as well as their own clinical observations was used as the reference standard. We have reported all aspects of the study recommended by STARD 2015 (S1 File).

### Sample size

We used the method proposed by Buderer, which allows to estimate the sample size required for clinically acceptable precision estimates of sensitivity and specificity, taking into consideration the prevalence of the disease in the study population(20):

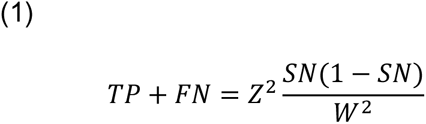

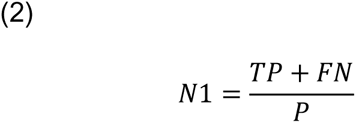

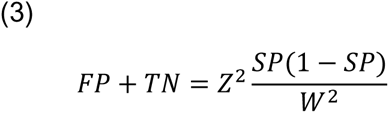

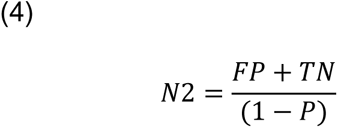

Where:

TP: true positives, TN: true negatives, TN: true negatives, FN: false negatives, SN: sensitivity, SP: specificity, Z: the standard normal, P: prevalence of the disease in the studied population, W: the confidence estimates for the prevalence.

(1) renders the number with a positive test, (2) renders the sample size required for sensitivity, (3) renders the number with a negative test and (4) renders the sample size required for specificity. The final sample size is the largest of N1 and N2.

For this study, we used the .85% and 74% for the clinically relevant sensitivity and specificity, based on previous data (13). Assuming a prevalence estimate of 25% and a significance level of 0.05, 192 participants were required for this assessment.

### Statistical analysis

Scores in the written training assessment were presented as a percentage. We assessed whether there were any associations between age, education level, gender and score using a Pearson’s coefficient for age, and a single factor ANOVA test for gender and education level.

For the scabies validation assessment, sensitivity, specificity, PPV (positive predictive value) and NPV (negative predictive value) were calculated for the diagnosis of scabies by comparing the index test to the reference standard. For this, “Observer 1” was collectively represented by FW1-3, and “Observer 2”, collectively represented by FW4-9.

Two Cohen’s kappa coefficients were calculated in order to assess the inter-rater agreement between medical doctors, and the rate of agreement between fieldworkers (21).

We used Excel (version 1808, Microsoft) and RStudio, version 4.1.1, (R Core Team (2022). R: A language and environment for statistical computing. R Foundation for Statistical Computing, Vienna, Austria. URL https://www.R-project.org/.) to analyse the data.

### Ethics approval

The study protocol was approved by the Internal Scientific Committee and Institutional Review board from the Centro de Investigação em Saúde de Manhiça (Ref: CIBS-CISM/004/2021), Hospital Clinic of Barcelona Clinical Research Ethics Committee (Ref: HCB/2019/0938) and The Ethics Research Committee of WHO (Protocol ID: ERC.0003265).

## RESULTS

### Demographic characteristics of fieldworkers

62 fieldworkers completed the training. The median age of the fieldworkers was 26 years and 65% were men (Table 1). The majority of participants (81.7%) had completed grade 12 of high school.

**Table 1.** Demographic characteristics of fieldworkers.

|  | N |  |
| --- | --- | --- |
| **Median age (IQR)** | 62 | 26 (24-29) |
| **Sex** |  |  |
| Male | 40 | 65% |
| Female | 22 | 35% |
| **Level of education\*** |  |  |
| Basic | 4 | 7% |
| Medium | 51 | 82% |
| Advanced | 7 | 11% |

\***Basic:** completed grade 10 of high school, **Medium:** completed grade 12 of high school,
**Advanced:** completed University degree.

### Training

The overall median results in written assessments 1 and 2 respectively were 75% and 85% and ranged from 46% to 92% (Table 2). The fieldworkers performed best in the identification of scabies lesions, with mean scores of 79% and 90% respectively. They had slightly lower scores in the scabies lesion count and the identification of infected scabies lesions. Whilst the scores were higher in all sections of assessment 2 compared to in assessment 1, the difference in scores was not statistically significant. There was no association between sex, age or level of education and scores (S1 Table).

**Table 2.**
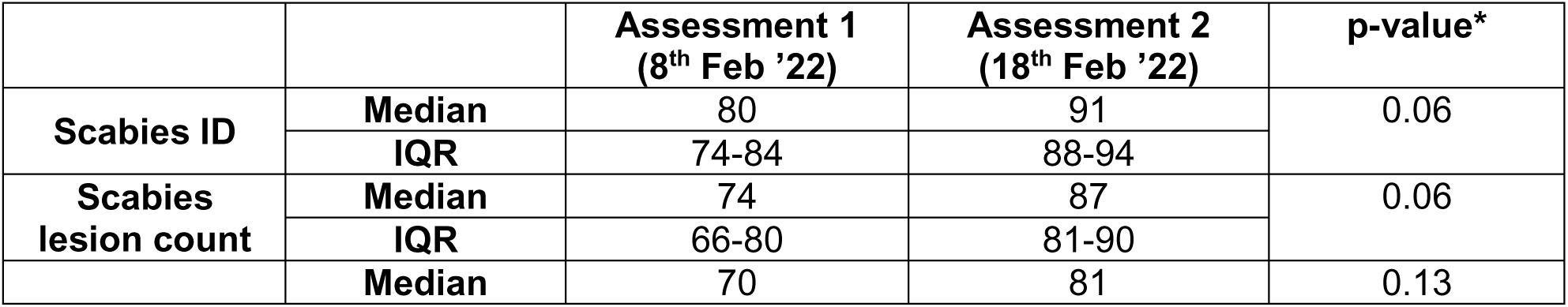
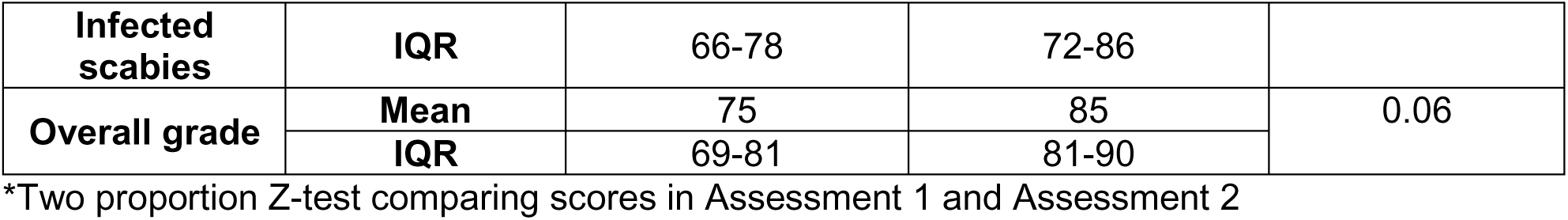
A comparison of fieldworker assessment results after initial training session and after the refreshment training session.

### Performance of fieldworkers in field settings

193 participants were selected from the 6 clusters reporting highest prevalence of scabies during visit 3 of the trial (1 month prior to the study). The median age of participants being examined for scabies was 16 years (IQR 6-29), and 53% were female (Table 3). According to the reference standard, 26% of these participants met the diagnostic criteria for scabies. 55% of those cases were severe, 39% moderate, 6% mild and 0% very mild. The majority (84%) of these cases were classified as clinical scabies.

**Table 3.** Demographic characteristics and spectrum of presenting symptoms in study participants.

|  |  |
| --- | --- |
| **Median age (IQR)** | 16 years (6-29) |
| **Sex, N (%)** |  |
| Male | 90 (47%) |
| Female | 103 (53%) |
| **Total Scabies** | 51 (26%) |
| **IACS criteria subcategory** |  |
| Clinical | 43 (22%) |
| Suspected | 8 (4%) |
| Not scabies | 142 (74%) |
| **Severity of scabies** |  |
| Very mild | 0 |
| Mild | 3 (6%) |
| Moderate | 20 (39%) |
| Severe | 28 |

Each of the nine fieldworkers who took part in the study assessed varying numbers of participants with varying proportions of true scabies cases, as indicated in Table 4.

**Table 4.** Number of positive and negative scabies assessments per fieldworker according to the reference standard and the fieldworker.

| Observer | Fieldworker (index) | Participants screened, N | Scabies (clinical or suspected), N |  | No scabies, N |  |
| --- | --- | --- | --- | --- | --- | --- |
|  |  |  | REF | FW | REF | FW |
| 1 | FW1 | 141 | 38 | 36 | 103 | 105 |
|  | FW2 | 33 | 0 | 1 | 33 | 32 |
|  | FW3 | 19 | 13 | 15 | 6 | 4 |
| 2 | FW4 | 26 | 0 | 2 | 26 | 24 |
|  | FW5 | 32 | 9 | 12 | 23 | 20 |
|  | FW6 | 33 | 0 | 0 | 33 | 33 |
|  | FW7 | 33 | 19 | 18 | 14 | 15 |
|  | FW8 | 33 | 15 | 16 | 18 | 17 |
|  | FW9 | 36 | 8 | 8 | 28 | 28 |

The sensitivity of fieldworker diagnosis compared to medical doctor diagnosis was 95.5% (95%CI 93.0 -99.9), and the specificity was 99.6% (95%CI 99.0-100) (Table 5). The sensitivity of the detection of severe scabies cases was lower at 64.3% (95%CI 50.9-74.9), and the mean specificity was 100% as fieldworkers were more likely to classify a severe case as moderate/mild than vice versa. However, for moderate cases the sensitivity and specificity remained similar. With respect to diagnosis of scabies (without consideration of severity or whether the case was clinical/suspected), the kappa coefficient indicated perfect agreement between the medical doctors, with a value of 1.00. For the fieldworkers, the kappa coefficient also indicated high levels of agreement, with a value of 0.95. Full 2×2 tables are available in S2 Table.

**Table 5.** Diagnostic accuracy of scabies diagnoses made by fieldworkers.

|  | Mean sensitivity (95%CI) | Mean specificity (95%CI) | Mean PPV (95%CI) | Mean NPV (95%CI) | MD kappa coefficient | FW kappa coefficient |
| --- | --- | --- | --- | --- | --- | --- |
| **Scabies (clinical or suspected)** | 95.5 (93.0-99.9) | 99.6 (99.0-100) | 99.0 (97.3-100) | 98.2 (97.3-100.0) | 1.00 | 0.95 |
| **Clinical** | 97.7 (88.4-98.5) | 98.7 (98.0-100) | 95.3 (94.2-99.8) | 99.3 (96.5-99.6) | 0.96 | 0.97 |
| **Severe** | 64.3 (50.9-74.9) | 100 (100-100) | 100 (100-100) | 94.4 (91.0-96.1) | 0.84 | 0.53 |
| **Moderate** | 95.7 (88.8-100) | 100 (100-100) | 100 (100-100) | 99.4 (98.7-100) | 0.90 | 0.86 |
| **Mild/very mild** | 100 (100-100) | 92.0 (89.3-94.7) | 16.22 (4.3-28.1) | 100 (100-100) | 0.80 | 0.35 |

## DISCUSSION

There is a significant gap in knowledge with respect to scabies prevalence in low and middle-income countries, particularly in SSA. Our reliance on highly trained medical doctors and light microscopy for analysis of skin scraping samples means that it is often not feasible to conduct prevalence surveys. Existing research has indicated that nurses or other health professionals are capable of diagnosing scabies with a moderate-high level of accuracy. The results of this study suggest that individuals without any medical training may also be able to screen individuals for scabies with a similar degree of accuracy as medical doctors after a short, focal training. Given the global shortage of healthcare workers and costs associated with more specialized staff, this may provide a cost-effective alternative method of assessing scabies prevalence.

The results of the training assessments showed that fieldworkers with only secondary education were able to achieve high scores after a short period of training. Our results suggested that this is true regardless of sex, age or level of education (beyond secondary school). When evaluating the accuracy of scabies (suspected or clinical) assessments in the field, our results suggested that diagnoses made by minimally trained fieldworkers compared to those by highly qualified medical doctors, were highly sensitive and specific when considering diagnosis of scabies (either suspected or clinical). When considering the assessment of severity, fieldworker assessments were less sensitive, particularly for severe scabies; there was a tendency to falsely assess severe scabies cases as milder. There also appeared to be less agreement between medical doctors and between fieldworkers with respect to grading the severity of disease. The values of diagnostic accuracy were higher than expected and higher than observed in similar studies. This was most likely due to limitations in the choice of reference standard, as detailed below.

The main limitation of this study was that the reference standard used did not reflect the gold standard and therefore the values are not representative of the true diagnostic accuracy. The gold standard for scabies diagnosis typically involves examination of the full body, often using dermoscopy for magnification of suspected lesions and burrows, and microscopy of skin scrapings to confirm the presence of mites or mite products. In this study, however, medical doctors examined only exposed skin, and did not take any samples for laboratory confirmation. Furthermore, due to the need of a translator in this study, the history of itch and contact history were shared between both fieldworkers and medical doctors. Although the reference standard used in our study is not perfect, it is similar to the methods typically used within the preventative medicine team in the study area. The proportion of severe cases in the study sample according to the reference standard is much higher than in other similar prevalence surveys, possibly suggesting that there were more mild cases of scabies that were not identified by the medical doctors (13,22). This may also be a factor contributing to the high diagnostic accuracy we observed.

## CONCLUSIONS

Despite the limitations of the study, the results do suggest that minimally trained workers with secondary school education are capable of diagnosing scabies to a similar degree of accuracy to trained medical doctors. Given the paucity of scabies prevalence data at present, and the limited resources available to support data collection, this may provide a valuable tool for rapid assessment of scabies prevalence where more specialized staff are not available. MDA programmes used for parasitic diseases including malaria, lymphatic filariasis or scabies may offer an opportunity for active case detection and monitoring of scabies. Further evaluations would be needed to better understand the true diagnostic accuracy of these assessments, how factors such as prevalence affect accuracy, and the contexts in which they are appropriate.

## Data Availability

All data produced are available online at: https://dataverse.csuc.cat/dataset.xhtml?persistentId=doi:10.34810/data1100

## STATEMENTS

### Author contributions

Conceptualisation: JFA, RR, DE, CCh

Data curation: JFA, EE

Formal analysis: JFA

Investigation: JFA, VL, HM, AH, AM, AX, FM

Methodology: JFA, DE, CCh

Supervision: RR, FS, CCh

Writing - original draft: JFA, CCh

Writing - review & editing: all authors contributed, reviewed and approved the last draft.

## Acknowledgements

The authors would like to thank all the participants and study team that took part in this study.

## Data sharing statement

All data and code relevant to this article are publicly available in the dataverse of the University of Barcelona: https://dataverse.csuc.cat/dataset.xhtml?persistentId=doi:10.34810/data1100

## Supplemental material

**S1 File. STARD Checklist**

**S1 Table. Associations between assessment grade, and age, sex, education level.**

**S2 Table. Diagnostic accuracy 2×2 table of sensitivity, specificity, PPV and NPV values for each observer**

