## Supplementary material for "Minimal training of fieldworkers in resource-poor settings for rapid assessment of scabies prevalence: a diagnostic accuracy study in Mopeia, Mozambique": S1 Table

**S1 Table. Associations between assessment grade, and age, sex, education level.**

| Variable | | N | Mean grade | Statistical test |
|---|---|---|---|---|
| Sex | Female | 22 | 82.39 | 0,20* |
| | Male | 36 | 83.76 | |
| Level of education | Basic | 4 | 88.33 | 0.09* |
| | Medium | 46 | 85.20 | |
| | Advanced | 8 | 80.67 | |
| Age | | 58 | | -0.17367** |

*p-value (Single-Factor ANOVA test)

**Pearson’s coefficient
