## Supplementary material for "Minimal training of fieldworkers in resource-poor settings for rapid assessment of scabies prevalence: a diagnostic accuracy study in Mopeia, Mozambique": S2 Table

S2 Table. Diagnostic accuracy for each index test.

| | TP (N) | FP (N) | TN (N) | FN (N) | Total (N) | Sensitivity | Specificity | PPV | NPV |
|---|---|---|---|---|---|---|---|---|---|
| **Observer 1** | 51 | 1 | 141 | 0 | 193 | 100 | 99.30 | 98.08 | 100 |
| **Observer 2** | 51 | 0 | 137 | 5 | 193 | 91.07 | 100 | 100 | 96.48 |
| **Total** | 102 | 1 | 278 | 5 | 386 | 95.53 | 99.64 | 99.03 | 98.23 |

TP: true positives, FP: false positives, FN: false negatives, TN: true negatives, PPV: positive predictive value, NPV: negative predictive value
